# High throughput sequencing based detection of SARS-CoV-2 prevailing in wastewater of Pune, West India

**DOI:** 10.1101/2021.06.08.21258563

**Authors:** Tanmay Dharmadhikari, Vinay Rajput, Rakeshkumar Yadav, Radhika Boaragaonkar, Dayanand Panse, Sanjay Kamble, Syed Dastager, Mahesh Dharne

**Affiliations:** National Collection of Industrial Microorganisms (NCIM), Biochemical Sciences Division, CSIR-National Chemical Laboratory, Pune, India; Academy of Scientific and Innovative Research (AcSIR), Ghaziabad, India; Ecosan Services Foundation (ESF), Pune, India; Chemical Engineering and Process Development Division, CSIR-National Chemical Laboratory, Pune, India

**Keywords:** wastewater, epidemiology, nanopore sequencing, metagenomics, SARS-CoV-2, ARTIC protocol

## Abstract

Given a large number of SARS-CoV-2 infected individuals, clinical detection has proved challenging. The wastewater-based epidemiological paradigm would cover the clinically escaped asymptomatic individuals owing to the faecal shedding of the virus. We hypothesised using wastewater as a valuable resource for analysing SARS-CoV-2 mutations circulating in the wastewater of Pune region (Maharashtra; India), one of the most affected during the covid-19 pandemic. We conducted a case study in open wastewater drains from December 2020-March 2021 to assess the presence of SARS-CoV-2 nucleic acid and further detect mutations using ARTIC protocol of MinION sequencing. The analysis revealed 108 mutations across six samples categorised into 40 types of mutations. We report the occurrence of mutations associated with B.1.617 lineage in March-2021 samples, simultaneously also reported as a Variant of Concern (VoC) responsible for the rapid increase in infections. The study also revealed four mutations; S:N801, S:C480R, NSP14:C279F and NSP3:L550del not currently reported from wastewater or clinical data in India but reported in the world. Further, a novel mutation NSP13:G206F mapping to NSP13 region was observed from wastewater. Notably, S:P1140del mutation was observed in December 2020 samples while it was reported in February 2021 from clinical data, indicating the instrumentality of wastewater data in early detection. This is the first study in India to conclude that wastewater-based epidemiology to identify mutations associated with SARS-CoV-2 virus from wastewater as an early warning indicator system.

## 1. Introduction

The respiratory distress virus, Severe Acute Respiratory Syndrome – Corona Virus – 2 (SARS-CoV-2), has unprecedented effects on human life and the healthcare system worldwide. The findings of Tang et al. 2020 revealed the high viral load in the faecal matter of infected individual, irrespective of the individuals showing any symptoms. The wastewater containing viral load from infected individuals would enter the wastewater system of well-planned sewage treatment plants or directly into the river system as untreated wastewater, raising concerns worldwide. The diagnostics are limited to the clinical context, and the wastewater system was hypothesised to gain insight into the thorough infection dynamics of the population. The Wastewater-based Epidemiological (WBE) approach would provide a comprehensive depiction of infection dynamics in the population by enabling asymptomatic individuals to be included, who would otherwise escape the clinical settings. The WBE approach was previously employed to identify illicit drug use and specific infective agents like SARS and Polio (Zuccato et al 2005, Heijnen and Medema 2009, Lago et al. 2003). This led towards the foundation of the WBE tool quickly for a better understanding of SARS-CoV-2 spread worldwide.

It was crucial to evaluate the current wastewater viral concentration protocols to optimize viral detection of the novel virus, and the work started promptly. Warish et al. 2020 provided an evidence-based protocol for concentrating Murine Hepatitis Virus, a positive sense single-stranded enveloped virus, as a surrogate for SARS-CoV-2, from wastewater using seven methods and the MCE protocol showed the highest recovery. Simultaneously, the work for isolating and concentrating SARS-CoV-2 from wastewater was also started worldwide. The statistical model-based evidence suggested by Peccia et al. 2020 provided insight into the correlation of the fluctuations observed in the number of infected individuals and viral load present in the wastewater. Detection of SARS-CoV-2 from the wastewater was also carried out worldwide to indicate the viral presence. Wastewater is the metagenomic landscape with various organisms; hence, detecting specific viral nucleic acids posed a challenge (Che et al. 2019). The MinION sequencer from Oxford Nanopore Technologies can be very useful in such scenarios as the total genomic material from the sample can be sequenced to identify the potential candidate (Che et al. 2019). Here, the development of ARTIC protocol facilitated the study of the metagenomic landscape of SARS-CoV-2 from wastewater utilizing amplicon sequencing to obtain amplified whole-genome fragments and analyse the mutations (Tyson et al. 2020, Josh Quick 2020).

Multiple mutations of SARS-CoV-2 were observed worldwide and raised concerns about the effectiveness of treatment and vaccines. Particular mutants were speculated for the increased infection spread, such as the rapid spread of the B.1.617 lineage variant by mediating the increment of viral entry into certain cell lines (Hoffmann et al. 2021). Studies were also performed for the effectiveness of vaccine candidates among the Variants of Concern (VOC), such as the effectiveness of BBV152 (Covaxin) being able to generate neutralising serological response against B.1.617 lineage (Yadav et al. 2021). The tracking of genomic variants from wastewater was assumed essential to understand the spread. The phylogenetic assessment of SARS-CoV-2 from wastewater was carried out by Nemudryi et al. 2020 using a long-read sequencing platform. Further, genomic variants were studied using Next Generation Sequencing (NGS) platforms by Agrawal et al. 2021 in Germany, Landgraff et al. 2021 in Canada, Wilton et al. 2021 in London, Crits-Christoph et al. 2021 in California, Jahn et al. 2021 in Switzerland and others. The studies were essential to analyse the regionally prevalent mutations in circulation, aiding the assumption of Variants of Concerns as causal elements in rising cases in the region.

Presently India is one of the worst affected countries globally, and the Pune region in the state of Maharashtra recorded one of the highest CoViD19 infections (The Times of India: Maharashtra reports the highest single-day spike of 63,729 Covid-19 cases). It was necessary to evaluate the wastewater from Pune city to understand the infection dynamics and focus on the mutations circulating in the population. However, no studies are currently being recorded in India, allowing the mutation analysis of SARS-CoV-2 from wastewater. To emphasise the importance of the mutation study from wastewater, we present the first study in India for the amplicon-based metagenomic landscape of SARS-CoV-2 in the wastewater of the Pune region.

We hypothesised that wastewater in Pune, being a highly affected region, would demonstrate SARS-CoV-2 RNA presence, which eventually could be employed to analyse the genomic mutations. Our goal was to examine the presence of SARS-CoV-2 RNA in the wastewater streams and employ the NGS platform of the MinION sequencer to identify mutations. The study could provide essential information regarding the mutations circulating in the community while also examining the potential source of wastewater as an early warning system.

## 2. Material and Methods

**Figure 1.**
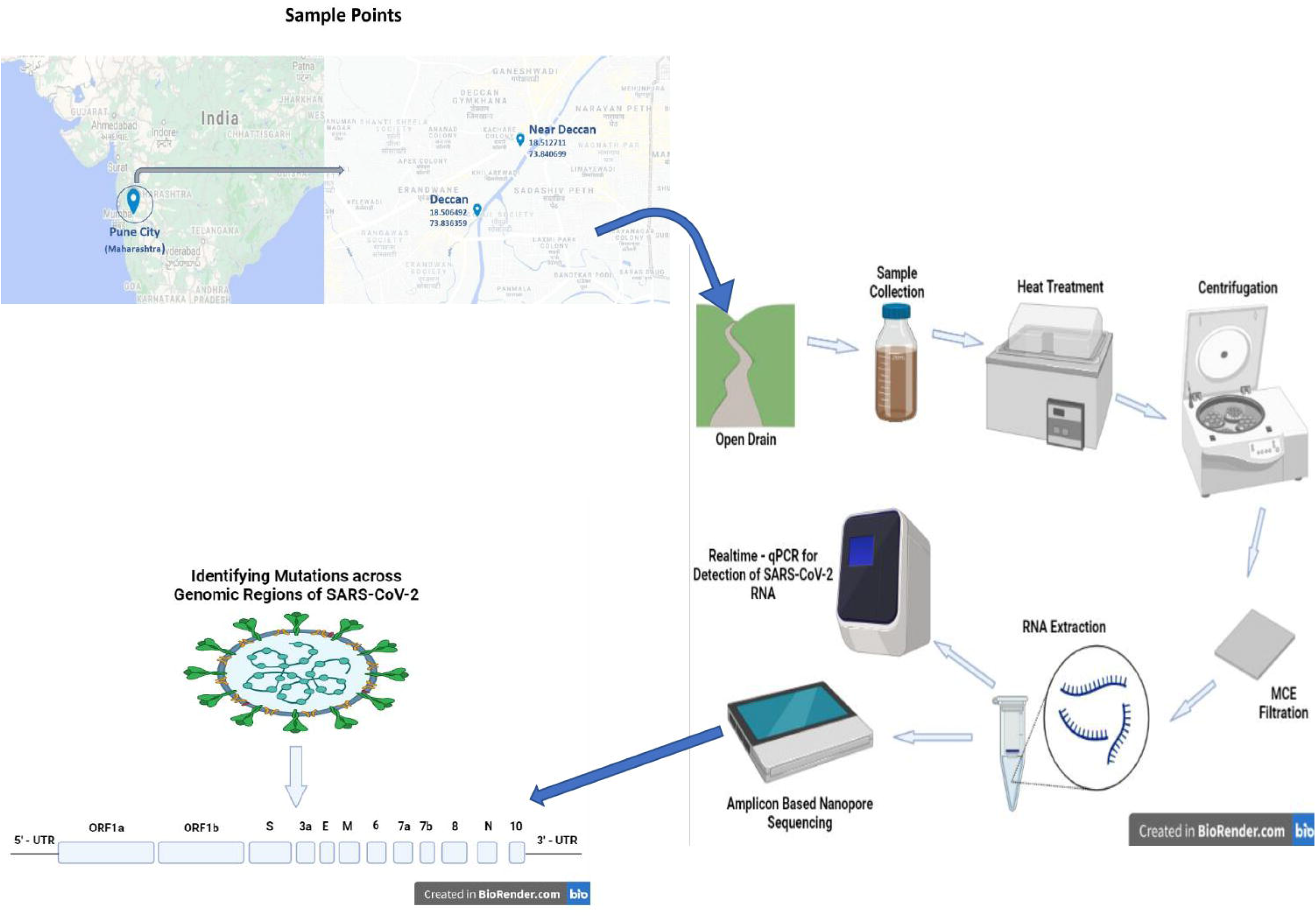
Process includes sample collection from two sites. The sample were processed and genomic mutations were analysed.

### 2.1 Sample Collection and Processing

The Sampling site Deccan (18.506492,73.836359; Kothrud Basin) and Near Deccan (18.512711,73.840699; Prabhat Road Basin) are open wastewater drains entering the Mutha river near the sample collection site. The wastewater samples WW9, WW10 and WWP (WW1, WW2 and WW3 except for WW4) were collected from the Deccan site and WW8, WW10, WW12 and WW4 (from WWP) from the Near Deccan site. A total of 12 composite samples were collected as 1 litre-1hour grab sample between morning 09:00 am to 10:00 am in a sterile plastic container (Himedia Solution Bottles - TCP040-1×12NO) throughout December 2020 to March 2021 (Rimoldi et al. 2020). The sample bottles were thoroughly cleaned from outside by 70% alcohol and 1% hypochlorite solution and transported to the laboratory at 4°C. The container with wastewater samples was kept in the water bath for 60 minutes at 60°C for heat inactivation (Wang et al. 2020). After the heat treatment, the bottles were allowed to cool down to room temperature and were immediately processed. The permissions regarding sample collection and processing were obtained from Pune Municipal Corporation, and Institutional Biosafety and Ethical Committee.

### 2.2 Virus Concentration

An aliquot of 200ml was transferred into a sterile Fluorinated HDPE Bottle (Capacity 250: Tarson; 584230) and centrifuged at 4500g for 10 min to settle down larger debris. The sample was then filtered through Whatman filter paper (Millipore; 1001-070-100/pk (Grade 1 Circles, 70mm Whatman)) using the vacuum filtration assembly (Tarson; Membrane Filter Holder - 47 mm-500ML) and Vacuum pump. The filtrate was transferred into a sterile glass flask, and MgCl_2_ was added. The sample was then filtered through a 0.45um Mixed Cellulose Ester filter (Millipore; MILLIPORE MEMBRANE FILTER, MIXED CELLULOSE ESTER (0.45 µm, 47 mm)). The MCE filter was immediately transferred to a bead beating tube from the RNA extraction kit. Contaminated glassware and plastic wares were decontaminated or disposed according to the institutional regulations.

### 2.3 RNA extraction and Realtime-qPCR

RNeasy Power Water Kit (Qiagen; 14700-50-NF) was used for RNA extraction following the instruction by the manufacturer. The RNA was eluted in 100 µl of RNase free water and stored at -80°C until further molecular process. The Real-time quantitative Polymerase Chain Reaction (RT-qPCR) was performed for the detection of SARS-CoV-2. The eluted RNA stored at -80 °C was thawed on ice. The SARS-CoV-2 specific ICMR validated kit TRUPCR® SARS-CoV-2 RT qPCR kit (V-3.2) (3B BlackBio Biotech India Limited; 3B306) was used for detection on Applied Biosystem ™ 7500 plus (Applied Biosystems). The threshold for cycle cut-off was set manually, and positive samples were detected. The SARS-CoV-2 positive RNA was employed further for the Oxford Nanopore Sequencing platform using ARTIC protocol (Tyson et al. 2020, Josh Quick 2020).

### 2.4 cDNA and Nanopore Library Preparation

According to manufacturer instructions, the Real-time-qPCR positive RNA from six samples was subjected to cDNA preparation using Maxima H minus Reverse Transcriptase Enzyme (Thermofisher; EP0752). The cDNA prepared was then purified using Agencourt Ampure XP beads (Beckman Coulter; A63881). The purified cDNA was further subjected to nCoV-2019 sequencing protocol v3 (LoCost) V.3, which uses two primer pool to amplify the fragments of SARS-CoV-2 whole genome present in sample (Tyson et al. 2020, Josh Quick 2020). The reverse-transcribed cDNA was amplified using Q5 High-Fidelity DNA Polymerase (New England Biolabs; M0491S), 5X Q5 Reaction Buffer (New England Biolabs; M0491S), dNTPs mix (New England Biolabs; N0447S) and primer pools 40 U/ul SARS-CoV-2 primers (Pool A & B) 100uM (ARCTIC) (New England Biolabs; GTR_066_COVID25). The amplified pools were mixed and purified using Agencourt Ampure XP beads (Beckman Coulter; A63881). End preparation and Barcoding was performed using Blunt/TA Ligase MasterMix (New England Biolabs; M0367L), NEBNext Ultra II End Repair/dA-Tailing Module (New England Biolabs; E7546L) and Native Barcoding Expansion 1-12 (PCR-free) (Oxford Nanopore Technologies; EXP-NBD104). The quantification was performed with Qubit Fluorometer (Invitrogen), and the 24ng library was loaded onto the flow cell. The barcoded samples were pooled together, and the run was set up on the MinION device (Oxford Nanopore Technologies). The sequencing was allowed to run for 24 hrs, and data was collected. The raw reads from the Nanopore sequencer were base-called using Guppy High Accuracy - dna_r9.4.1_450bps_hac.cfg, and further analysis was carried out using ARTIC Bioinformatic Pipeline with few required modifications (Tyson et al. 2020, Josh Quick 2020). All the sequences obtained were analysed with reference to the SARS-CoV-2 reference genome Wuhan-Hu-1 (NCBI Accession: MN908947). The GSAID database was utilised to obtain information regarding the reported mutation and was last accessed on 01st May 2021.

## 3. Results

The Ct values of the samples are provided in the supplementary Table 1. All the wastewater samples in the study collected between December 2020 through March 2021 consistently were positive for SARS-CoV-2 nucleic acid fragments. The cycle threshold values obtained have shown variability attributed to the changing infection dynamics. The amplicon sequenced SRA data is submitted to the NCBI database with accession number SRA:PRJNA728440. The analysis revealed several mutations in multiple genomic regions of SARS-CoV-2, including 3’UTR, ORF1a, ORF1b, Spike, ORF3a, ORF7a, M, ORF6, N, ORF8 and 3’UTR. In total, 108 mutations, categorised into 40 types based on nucleotide position, were detected in all the samples (details in Supplementary Table 3). We detected 15 mutations from WW8, 19 mutations from WW9, 17 mutations from WW10, 20 mutations from WW11, 23 mutations from WW12 and 13 mutations from WW-P. Notably, nine mutations in the Spike region (S: L452R, S:C480R, S: E484Q, S: D614G, S: P681R, S: N801, S: D950N, S: Q1071H, S: P1140) were observed in this study. The March-2021 samples showed L452R and E484Q mutations, while these mutations were absent in the sample collected from December-2020 to February-2021 (WW-P). We detected five novel mutations not reported from Indian clinical sequence data on Global Initiative on Sharing Avian flu Data (GISAID) (Shu and McCauley 2017). These mutations are as follows: 23964 AT>A (S: N801del), 4369 TG>T (NSP3:L550), 18875 C>T (NSP14:C279F), 16852/16853 GG>TT (NSP15:C206F), 23000 T>C (S:C480R).

**Table 1.** Mutations identified in the six samples collected from December 2020 throughout March 2021.

| 5'UTR | ORF | NSP | N | S | M | 3'UTR | Samples |
| --- | --- | --- | --- | --- | --- | --- | --- |
| 5'-UTR:210<br>5'-UTR:241 | ORF3a:S26L | NSP3:Y246Y<br>NSP12b:P314L |  | S:P1140del |  |  | Common for All<br>Samples |
|  |  | NSP13:M429I |  | S:L452R<br>S:E484Q |  | 3'UTR:28270 | WW8, WW9, WW10,<br>WW11, WW12 |
|  |  |  |  | S:D614G |  |  | WWP, WW8, WW9,<br>WW10, WW11 |
|  |  | NSP3:T749I<br>NSP6:T77A |  | S:Q1071H |  |  | WW8, WW9, WW10,<br>WW11 |
|  | ORF6:I33T<br>ORF7a:V82A |  |  |  |  |  | WW9, WW10, WW11 |
|  |  |  |  |  |  | 3'UTR:29742 | WW8, WW9, WW11 |
|  | ORF3a:E261*<br>ORF8:S97I | NSP3:H1630H<br>NSP10:H80H |  |  |  |  | WWP |
| 5'-UTR:75 |  | NSP3:L550del<br>NSP13:P77L<br>NSP13:G206F<br>NSP13:V484F<br>NSP14:C279F |  | S:C480R<br>S:D950N | M:V10A | 3'-UTR:21555<br>3'UTR:26493 | WW12 |
|  |  | NSP14:C279C |  |  |  |  | WWP, WW12 |
|  |  |  |  | S:N801 |  |  | WWP, WW10 |
|  |  | NSP3:P822L |  |  |  |  | WW11, WW12 |
|  |  |  | N:R203M |  |  | 3'-UTR:29700 | WW9 |
|  |  |  | N:D63G | S:P681R |  |  | WW11 |

## 4. Discussion

The wastewater-based epidemiological approach can predict the population’s infection dynamics (Peccia et al. 2020). However, an exact estimation of the infected individuals is currently unattainable. The study to estimate exact viral load using recovery of concentration protocols and sustainability of virus from the faecal source of infected individuals to the endpoint of sewage treatment plant has not been carried out. However, the WBE study can be applied to obtain comparative infection dynamics of a particular region to obtain information regarding the severity of affected regions (Peccia et al. 2020). Since the wastewater has shown consistent viral presence, as seen in samples taken from December 2020 through March 2021, it is essential to bring the public attention to the viral presence and create awareness. The study also provided instances where the mutations obtained from the wastewater sequenced data are either not reported in GISAID from India or, in case of novel mutation, not across the world.

The WBE study can provide us with information regarding mutations indicating genomic variants in the population (Hoffman et al. 2021, Agrawat et al. 2021, Landgraff et al. 2021, Wilton et al. 2021, Crits-Christoph et al. 2021 and Jahn et al. 2021). The clinical evaluation of variants in circulation, where asymptomatic can be overlooked, along with the time-consuming protocols of sequencing, together creates an arduous exercise. Maharashtra (India) has recorded very high cases of infection, and it has been raised concern as variants of B.1.617 lineage to be a causal factor (Indian Express: 20. Explained: B.1.617 variant and the Covid-19 surge in India). The present wastewater sequencing analysis revealed mutations L452R and E484Q associated with B.1.617 lineage in samples collected during March 2021, while the mutations were absent in samples collected from December 2020 to February 2021. The clinical sequencing data also observed higher infections with B.1.617 lineage from a similar period. Here, it can be observed that regular wastewater monitoring for identifying mutations can be a critical resource, as it can act as an early warning system. Hence, regular monitoring of wastewater is an essential criterion to observe mutations associated with concerned variants in circulation as the required results can be obtained from a smaller sample volume of wastewater than the more significant number of individuals.

The mutations S:P1140del was reported in late February 2021 from clinical data in India and earlier only from Africa-Egypt and North America (GISAID - Shu and McCauley 2017). However, the mutation, mapped to the Spike region, was present in all the samples collected from December 2020 to March 2021. This observation provides evidence of how wastewater sequencing data identify the mutations indicating possible variant in circulation before being observed through clinical data. WW12 sample was collected in the last week of March 2021 and has 12 mutations unique to the sample, in which NSP3:L550del, NSP14:C279F and S:C480R are not presently reported from India (GISAID - Shu and McCauley 2017). We also report a novel mutation NSP13:G206F (NSP13 region) detected in the WW12 sample. This occurrence of a novel mutation across the wastewater sample can be an instance of mutations observed before they are identified clinically. The WWP is the pooled sample from WW1, WW2, WW3 and WW4 from Deccan and Near Deccan site presenting clinically reported 13 mutations, and we also report the mutation S:N801 (Spike region), not presently reported from India (GISAID - Shu and McCauley 2017). WW9 has shown mutation N:R203M and 3’-UTR:29700, while WW11 has shown mutation S:P618R and N:D63G, present in samples collected during March 2021, which were absent in sample WWP taken before that. It can be predicted that the mentioned mutations prevailed in wastewater from March 2021 and might have been absent before. These mutations have not been reported from India, while they were reported in other countries (GISAID - Shu and McCauley 2017). This detection of mutation from wastewater sample provides an instance where clinically neglected mutations can be observed in wastewater sample, providing thorough information for mutations present in circulation.

Studies have observed that variants of the SARS-CoV-2 virus showed distinct infectivity effects (Hoffmann et al. 2021). In order to understand the circulating mutations of SARS-CoV-2, the wastewater system can provide thorough information regarding the mutations and analysing variants. The methods used in the experiment followed the protocols of Ahmed et al. 2020 for viral concentration. The virus concentration using MCE electronegative filtration provides a mean recovery of 65.7% ± 23.8 (Mean ± SD of % recovery) of the viral particles from the sample (Ahmed et al. 2020). Regular surveillance can increase the possibility of concentrating virus to observe prominent mutations. The studies conducted in India have reported the viral presence across the STPs or wastewater. However, no studies have yet revealed NGS platform sequencing to identify the mutations in circulation from wastewater (Srivastava et al. 2021, Chakraborty et al. 2021, Hemalatha et al. 2021, Arora et al. 2020 and Kumar et al. 2020). Our study is the first to report mutations through WBE, provide insight into circulating variants, and report novel mutations.

## 5. Conclusion

Wastewater can be considered a vital source to understand the mutations in circulation and understand the infection dynamics. The prevalence of particular mutations in circulation and clinically unreported mutations have been reported in this wastewater study of SARS-CoV-2, providing conclusive evidence for the potential utilization of wastewater. We also report novel mutations such as NSP13:G206F (NSP13), which conclude the capability of wastewater sequencing data to provide mutations in circulation before they are observed clinically. Regular monitoring of wastewater system for analysing mutations can act as an early warning system to understand the community infection dynamics.

## Data Availability

The amplicon sequenced SRA data is submitted to the NCBI database with accession number SRA:PRJNA728440.

## Author Contributions

All authors have read and review the manuscript for submission. TD: performing experiments, writing original draft and art design; RKY: performing experiments, editing and review; VR: bioinformatic analysis, review and editing; RB: review; DP: review; SK: study plan and review; SD: review and funding; MD: editing, review, funding and corresponding author

## Acknowledgements

Authors are indebted to the Director, CSIR-NCL for providing facilities, infrastructure and support. Authors thank Pune Municipal Corporation for providing the required permissions and partnering organisation Ecosan Services Foundation for their support in obtaining permissions and sampling. TD, VR and RKY acknowledge SERB and University Grant Commission (UGC), India respectively for fellowship support. Authors also extend acknowledgement to BioRender.com for creation of images. Authors thank Mr. Nitya Jacob, India Coordinator, Sustainable Sanitation Alliance (SuSanA) India Chapter for providing technical guidance in the study.

