## Supplementary Data for "High throughput sequencing based detection of SARS-CoV-2 prevailing in wastewater of Pune, West India"

Supplementary Table 1: Sample collection sites, sample collection dates and Realtime-qPCR results

| **Sample ID** | **Site** | **Date of Collection** | **Ct values** | **Result** |
| --- | --- | --- | --- | --- |
| WW1 | Deccan | 23-12-2020 | 29.9 | Positive |
| WW2 | Deccan | 26-12-2020 | 32.4 | Positive |
| WW3 | Deccan | 19-02-2021 | 31 | Positive |
| WW4 | Near Deccan | 22-02-2021 | 34 | Positive |
| WW5 | Deccan | 02-03-2021 | 33.4 | Positive |
| WW6 | Near Deccan | 02-03-2021 | 32.3 | Positive |
| WW7 | Deccan | 10-03-2021 | 33.8 | Positive |
| WW8 | Near Deccan | 10-03-2021 | 30.4 | Positive |
| WW9 | Deccan | 16-03-2021 | 32.6 | Positive |
| WW10 | Near Deccan | 16-03-2021 | 31.8 | Positive |
| WW11 | Deccan | 30-03-2021 | 31.9 | Positive |
| WW12 | Near Deccan | 30-03-2021 | 33 | Positive |

Supplementary Table 3: Mutations detected using ARTIC Bioinformatics Pipeline. (GISAID database last accessed on 01.05.2021)

|  | **Nucleotides** | | |  | **Gene** | **Status** | **Coverage** | | **Sample ID** |
| --- | --- | --- | --- | --- | --- | --- | --- | --- | --- |
| **#CHROM** | **POS** | **REF** | **ALT** | **Amino Acid Changes** | **Position** |  | **Total** | **Variant** |  |
| MN908947.3 | 75 | C | T | 5'-UTR:75 | 5'-UTR | Extragenic | 301x | 145x | WW12 |
| MN908947.3 | 210 | G | T | 5’-UTR:210 | 5'-UTR | Extragenic | 69x  558x 694x 823x 1248x 313x | 34x  337x 456x 474x 1014x 248x | WWP  WW8 WW9 WW10 WW11 WW12 |
| MN908947.3 | 241 | C | T | 5’-UTR:241 | 5'-UTR | Extragenic | 69x  558x 694x 823x 1248x 313x | 59x  436x 527x 649x 969x 249x | WWP  WW8 WW9 WW10 WW11 WW12 |
| MN908947.3 | 3457 | C | T | NSP3:Y246Y | NSP3 | Nonsynonymous | 80x  89x 108x 152x 555x 86x | 51x  59x 82x 87x 338x 85x | WWP  WW8 WW9 WW10 WW11 WW12 |
| MN908947.3 | 4369 | TG | T | NSP3:L550del | NSP3 | Not Reported in India | 500x | 291x | WW12 |
| MN908947.3 | 4965 | C | T | NSP3:T749I | NSP3 | Reported in India and in World | 1054x 1451x 3807x 4788x | 602x 1030x 2133x 2723x | WW8 WW9 WW10 WW11 |
| MN908947.3 | 5184 | C | T | NSP3:P822L | NSP3 | Reported in India and in World | 465x 374x | 380x 339x | WW11 WW12 |
| MN908947.3 | 7609 | C | T | NSP3:H1630H | NSP3 | Nonsynonymous | 70x | 41x | WWP |
| MN908947.3 | 11201 | A | G | NSP6:T77A | NSP6 | Reported in India and in World | 766x 754x 1048x 1344x | 335x 453x 476x 614x | WW8 WW9 WW10 WW11 |
| MN908947.3 | 13264 | C | T | NSP10:H80H | NSP10 | Nonsynonymous | 1306x | 774x | WWP |
| MN908947.3 | 14408 | C | T | NSP12b:P314L | NSP12 | Reported in India and in World | 684x  1250x 1176x 3212x 2745x 34x | 543x  1003x 974x 2520x 2148x 24x | WWP  WW8 WW9 WW10 WW11 WW12 |
| MN908947.3 | 16466 | C | T | NSP13:P77L | NSP13 | Reported in India and in World | 11357x | 5299x | WW12 |
| MN908947.3 | 16852 | GG | TT | NSP13:G206F | NSP13 | Novel Mutation | 558x | 395x | WW12 |
| MN908947.3 | 16853 |  |  |  |  |  | 558x | 391x | WW12 |
| MN908947.3 | 17523 | G | T | NSP13:M429I | NSP13 | Reported in India and in World | 397x 406x 986x 488x 73x | 228x 202x 436x 229x 53x | WW8 WW9 WW10 WW11 WW12 |
| MN908947.3 | 17686 | G | T | NSP13:V484F | NSP13 | Reported in India and in World | 73x | 54x | WW12 |
| MN908947.3 | 18875 | G | T | NSP14:C279F | NSP14 | Not reported in India | 271x | 98x | WW12 |
| MN908947.3 | 18877 | C | T | NSP14:C279C | NSP14 | Nonsynonymous | 271x. 1205x | 142x, 890x | WW12, WWP |
| MN908947.3 | 21555 | A | G | 3’-UTR:21555 | 3’-UTR | Extragenic | 334x | 174x | WW12 |
| MN908947.3 | 22917 | T | G | S:L452R | Spike | Reported in India and in World | 2809x 1531x 7579x 111x 18x | 205x 1152x 4659x 100x 16x | WW8 WW9 WW10 WW11 WW12 |
| MN908947.3 | 23000 | T | C | S:C480R | Spike | Not reported in India | 18x | 5x | WW12 |
| MN908947.3 | 23012 | G | C | S:E484Q | Spike | Reported in India and in World | 2815x 1536x 7581x 111x 18x | 2156x 1108x 4826x 88x 15x | WW8 WW9 WW10 WW11 WW12 |
| MN908947.3 | 23403 | A | G | S:D614G | Spike | Reported in India and in World | 1868x  10101x 13250x 10101x 6533x | 1565x  8455x 11255x 8458x 5398x | WWP  WW8 WW9 WW10 WW11 |
| MN908947.3 | 23604 | C | G | S:P681R | Spike | Reported in India and in World | 401x | 371x | WW11 |
| MN908947.3 | 23964 | AT | A | S:N801 | Spike | Not reported in India | 428x, 137x | 141x, 51x | WW10, WWP |
| MN908947.3 | 24410 | G | A | S:D950N | Spike | Reported in India and in World | 384x | 185x | WW12 |
| MN908947.3 | 24775 | A | T | S:Q1071H | Spike | Reported in India and in World | 7228x 6641x 4542x 8246x | 4036x 4082x 2001x 5241x | WW8 WW9 WW10 WW11 |
| MN908947.3 | 24981 | CT | C | S:P1140del | Spike | Reported in India and in World | 2167x  7165x 6561x 4484x 8241x 1940x | 638x  2158x 1945x 1355x 2506x 581x | WWP  WW8 WW9 WW10 WW11 WW12 |
| MN908947.3 | 25469 | C | T | ORF3a:S26L | ORF3a | Reported in India and in World | 1273x  2778x 2805x 4971x 3432x 91x | 726x  1533x 1792x 2049x 2463x 69x | WWP  WW8 WW9 WW10 WW11 WW12 |
| MN908947.3 | 26173 | G | T | ORF3a:E261* | ORF3a | Reported in India and in World | 237x | 200x | WWP |
| MN908947.3 | 26493 | GT | G | 3’UTR:26493 | 3’-UTR | Extragenic | 79x | 14x | WW12 |
| MN908947.3 | 26551 | T | C | M:V10A | M | Reported in India and in World | 66x | 23x | WW12 |
| MN908947.3 | 27299 | T | C | ORF6:I33T | ORF6 | Reported in India and in World | 1598x 4390x 4249x | 648x 2073x 2071x | WW9 WW10 WW11 |
| MN908947.3 | 27638 | T | C | ORF7a:V82A | ORF7a | Reported in India and in World | 22x 22x 112x | 16x 12x 84x | WW9 WW10 WW11 |
| MN908947.3 | 28183 | G | T | ORF8:S97I | ORF8 | Reported in India and in World | 1849x | 886x | WWP |
| MN908947.3 | 28270 | TA | T | 3’UTR:28270 | 3'-UTR | Extragenic | 5026x 4199x 3249x 3108x 3541x | 3447x 2934x 1884x 2338x 2816x | WW8 WW9 WW10 WW11 WW12 |
| MN908947.3 | 28461 | A | G | N:D63G | N | Reported in India and in World | 124x | 79x | WW11 |
| MN908947.3 | 28881 | G | T | N:R203M | N | Reported in India and in World | 33x | 26x | WW9 |
| MN908947.3 | 29700 | A | G | 3’-UTR:29700 | 3’-UTR | Extragenic | 33x | 13x | WW9 |
| MN908947.3 | 29742 | G | T | 3’UTR:29742 | 3'-UTR | Extragenic | 30x 33x 716x | 22x 25x 507x | WW8 WW9 WW11 |
